# Combining symbolic regression with the Cox proportional hazards model improves prediction of heart failure deaths

**DOI:** 10.1101/2021.01.15.21249874

**Authors:** Casper Wilstup, Chris Cave

**Affiliations:** Abzu, Gammel Lundtoftevej 3B, 2800 Kgs. Lyngby, Denmark

## Abstract

Heart failure is a clinical syndrome characterised by a reduced ability of the heart to pump blood. Patients with heart failure have a high mortality rate, and physicians need reliable prognostic predictions to make informed decisions about the appropriate application of devices, transplantation, medications, and palliative care. In this study, we demonstrate that combining symbolic regression with the Cox proportional hazards model improves the ability to predict death due to heart failure compared to using the Cox proportional hazards model alone.

We used a newly invented symbolic regression method called the QLat-tice to analyse a data set of medical records for 299 Pakistani patients diagnosed with heart failure. The QLattice identified a minimal set of mathematical transformations of the available covariates, which we then used in a Cox model to predict survival.

An exponential function of age, the inverse of ejection fraction, and the inverse of serum creatinine were identified as the best risk factors for predicting heart failure deaths. A Cox model fitted on these transformed covariates had improved predictive performance compared with a Cox model on the same covariates without mathematical transformations.

Symbolic regression is a way to find transformations of covariates from patients’ medical records which can improve the performance of survival regression models. At the same time, these simple functions are intuitive and easy to apply in clinical settings. The direct interpretability of the simple forms may help researchers gain new insights into the actual causal pathways leading to deaths.

## Background

Heart failure (HF) is a clinical syndrome characterised by a reduction in the ability of the heart to pump or fill with blood. HF can be defined physiologically as an inadequate cardiac output to meet metabolic demands, often manifesting as increased left ventricular filling pressure[13]. Among the causes of HF are coronary heart disease, hypertension, diabetes, obesity, and smoking[15]. HF affects at least 26 million people globally and has a high mortality rate[11].

Various methods have been developed to estimate the risk of death for patients with HF. Well-known models include the ADHERE model[9] and the Seattle Heart Failure Model[10]. Although these models are accurate, they are unintuitive and rely on extensive medical records, making them hard to apply in a clinical setting.

Ahmad et al. published a study of 299 patients with HF admitted to Faisalabad Institute of Cardiology or Allied Hospital Faisalabad, Punjab, Pakistan[3]. In their study, Ahmad et al. used a Cox model to predict survival of patients using much fewer covariates than the Seattle Heart Failure Model. The data set was made freely available by Ahmad et al., and it has subsequently been used in additional analyses using both survival models[17] and machine learning techniques[5].

### Cox proportional hazards model

The Cox model[6] is widely used to predict survival in health science. When using the method, the researcher needs to choose which risk factors to include in the model, and the final model performance is obviously dependent on the appropriate choice of risk factors.

The general form of the Cox model is[6]:

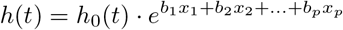

where

- *t* is the survival time
- *h*(*t*) is the hazard function determined by a set of p covariates (*x*_1_, *x*_2_, …, *x*_*p*_)
- (*b*_1_, *b*_2_, …, *b*_*p*_) are the coefficients which measure the impact of the covariates
- the term *h*_0_(*t*) is the baseline hazard, i.e. the hazard if all the *x*_*i*_ are equal to zero

This can also be written as:

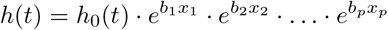

If we let *h*(*t*) and *h*^*/*^(*t*) be the hazard function for two individuals that differ only in *x*_1_ with a difference *δx*, their relative hazard is:

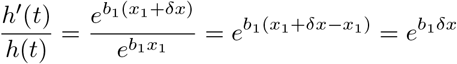

This shows an important limitation of Cox models: A unit change in a covariate will have the same effect on the hazard regardless of the origin of the change. For example: a change in *x*_1_ of 0.1 will have the same effect on survival whether from 1.0 to 1.1 or from 5.0 to 5.1.

### Mathematical transformations

In this paper, the term *mathematical transformation*, sometimes just *transformation*, means to apply a mathematical function to a covariate: *x*_*T*_ = *f* (*x*) for some function *f*. Perhaps the most widely used mathematical transformation is log-transformation: *x*_*T*_ = *log*(*x*). But other typical transformations include 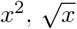, and *e*^*x*^.

When fitting a Cox model using mathematically transformed covariates, we have:

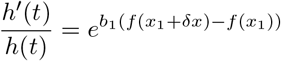

which is clearly no longer independent of the origin of *x*_1_ unless *f* is a linear function. Thus the effect of the transformation is to overcome this specific limitation in the generality of Cox models.

It is reasonable to hypothesise that well-chosen transformations of the available covariates will improve the predictive power of Cox models.

In this study, we demonstrate that this is indeed the case, and that a useful method to identify an appropriate mathematical transformation is to use symbolic regression.

### Symbolic regression

Symbolic regression is a machine learning method that attempts to explain some *Y* in terms of some *X* using a mathematical expression composed of a set of basic functions. However, the search space of possible expressions grows exponentially with the length of the expression, which makes a direct search infeasible. Traditionally, genetic programming has been used to search this space selectively[12, 16, 7]. Recent approaches have been more physics-inspired[14].

The QLattice is a symbolic regressor inspired by quantum field theory[1]. The QLattice runs on a dedicated high-performance computing cluster and models the list of possible mathematical expressions—in principle: infinite— that could link *Y* to *X* as a superposition of an infinite set of spatial paths. The QLattice searches this list of all mathematical expressions, including parameters, for the expressions that best model the output from the input. The result of the search is a list of expressions sorted by how well they match observations.

The QLattice can be used to generate either classification models or regression models. In this study, we only use it to generate classification models. Mathematically, this means that the QLattice will wrap each expression in a logistic function, thereby allowing the output to be interpreted as a probability. In other words: if *X* is an input vector, *f* (*X*) is the mathematical equation, and *Y* is the event we want to predict, then the QLattice will search for functions *f* such that the predictive power of:

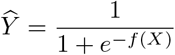

is maximised.

In this study, we first used symbolic regression to select and mathematically transform the available covariates from Ahmad et al. We subsequently fitted a Cox model to these transformed values.

To our knowledge, the combination of symbolic regression and Cox models has not previously been studied in research.

## Data

The data set used in this study consists of medical records of 299 HF patients admitted to the Faisalabad Institute of Cardiology and the Allied Hospital in Faisalabad (Punjab, Pakistan) between April and December 2015.

The data set contains 105 women and 194 men with left ventricular systolic dysfunction. The patients had HF in the classes III or IV of New York Heart Association (NYHA) classification scheme.

The patients were between 40 and 95 years old (mean 60.8) at the time of admission. The follow-up period was between 4 days and 285 days with a mean of 130 days.

The data set contains the following potential risk factors: age, serum sodium, serum creatinine, gender, smoking, blood pressure (BP), ejection fraction (EF), anaemia, platelets, and creatinine phosphokinase (CPK). Table 1 shows the summary statistics for the data.

Anemia in patients was assessed by their haematocrit level. Patients with haematocrit less than 36% (minimum normal level of haematocrit) were taken as anemic. Information on a patient’s smoking status and high blood pressure were taken from physician’s notes.

## Methods

In this study, we used the QLattice for two purposes: first, to select the three most important covariates from the data set for further modelling; and second, to find how these covariates can best be mathematically transformed to model risk of death. Finally, we fitted a Cox model on the transformed covariates.

### QLattice for feature selection

In the first step, we instructed the QLattice to search for a model that best described the probability of death given any three of the available covariates. The purpose of this search was to identify three important covariates that we then examined further.

**Table 1:** Summary statistics for the data set.

|  |  | Overall | Min | 25% | 50% | 75% | Max |
| --- | --- | --- | --- | --- | --- | --- | --- |
| n |  | 299 |  |  |  |  |  |
| TIME, mean (SD) |  | 130.3 (77.6) | 4 | 73 | 115 | 203 | 285 |
| Event, n (%) | 0 | 203 (67.9) |  |  |  |  |  |
|  | 1 | 96 (32.1) |  |  |  |  |  |
| Gender, n (%) | 0 | 105 (35.1) |  |  |  |  |  |
|  | 1 | 194 (64.9) |  |  |  |  |  |
| Smoking, n (%) | 0 | 203 (67.9) |  |  |  |  |  |
|  | 1 | 96 (32.1) |  |  |  |  |  |
| Diabetes, n (%) | 0 | 174 (58.2) |  |  |  |  |  |
|  | 1 | 125 (41.8) |  |  |  |  |  |
| BP, n (%) | 0 | 194 (64.9) |  |  |  |  |  |
|  | 1 | 105 (35.1) |  |  |  |  |  |
| Anaemia, n (%) | 0 | 170 (56.9) |  |  |  |  |  |
|  | 1 | 129 (43.1) |  |  |  |  |  |
| Age, mean (SD) |  | 60.8 (11.9) | 40 | 51 | 60 | 70 | 95 |
| EF, mean (SD) |  | 38.1 (11.8) | 14 | 30 | 38 | 45 | 80 |
| Sodium, mean (SD) |  | 136.6 (4.4) | 113 | 134 | 137 | 140 | 148 |
| Creatinine, mean (SD) |  | 1.4 (1.0) | 0.5 | 0.9 | 1.1 | 1.4 | 9.4 |
| Pletelets, mean (SD) |  | 263K (97K) | 25K | 212K | 262K | 303K | 850K |
| CPK, mean (SD) |  | 581.8 (970.3) | 23 | 116.5 | 250 | 582 | 7861 |

The user of the QLattice can choose the specific optimisation target to use when searching for models. We used the default optimisation target, which is to minimise the residual sum of squares.

When using the QLattice, it is possible to control the elementary mathematical functions from which the expressions are composed. For this search, we limited the QLattice to work with the following function set: 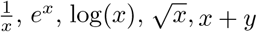, and *x · y*.

The QLattice searches through all the available input covariates in the data set, combines them into expressions using the elementary mathematical functions, and sorts them based on how well the resulting expression compares to the actual observations. Even given the limit of three inputs, the QLattice will search through millions of different expressions to find the best expression.

### QLattice for finding the best transformation of covariates

For each of the selected covariates, we used the QLattice to find the mathematical transformation that best predicted death events given that covariate alone.

Since this search only involved a single covariate, we limited the QLattice to work with the following unary function set: 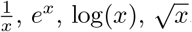.

### Cox Model

Finally, we fitted a Cox model with the output of the mathematically transformed covariates.

The resulting model was validated using concordance index (C-index). The C-index for a survival model can be thought of as the weighted average of the area under time-specific Receiver Operating Characteristic (ROC) curves. We also computed the time-specific ROC curve and area under the curve (AUC) for 285 day survival.

### Comparison Cox model

For comparison, we fitted a Cox model on the same covariates but *without* the identified mathematical transformations. The two models used the same covariates and only differed in the transformations. The models were compared using C-index, AUC (285 days), log-likelihood, and Akaike Information Criterion (AIC)[4].

All calculations were done in the programming language Python. For symbolic regression, we used QLattice version 1.4.6[1]. For Cox Modelling, we used lifelines version 0.25.7[8].

## Results

### Feature selection

The QLattice was given the opportunity to find the best model that uses at most three features out of the eleven. The model itself is unimportant as we want to model the survival function using Cox regression and not classify the data set into those that survived and those that died. The purpose of this step is to find which features carried the most signal.

The three features chosen by the QLattice when searching for symbolic forms were *ejection fraction, serum creatinine*, and *age*. This is consistent with other studies of the same data set using different feature selection methods[17, 5].

### Best transformation of Ejection Fraction

The best mathematical relation between ejection fraction and the probability of death was:

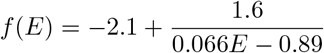

or, combined with the logistic wrapper:

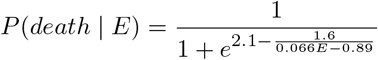

The probability of death was found to be closer associated with the inverse of the ejection fraction than with the ejection fraction directly.

### Best transformation of serum creatinine

The best mathematical relation between serum creatinine and the probability of death was:

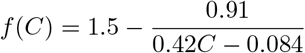

or, combined with the logistic wrapper:

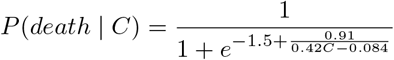

As with the ejection fraction, the probability of death is closer associated with the inverse of serum creatinine than to serum creatinine directly.

### Best transformation of age

The best mathematical relation between age and the probability of death was:

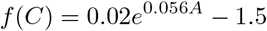

or, combined with the logistic wrapper:

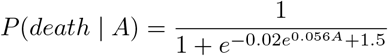

The probability of death grows exponentially with age, with a growth constant of 0.056.

### Cox model with transformed risk factors

Having determined that *e*^0.056*A*^, 1*/E*, and 1*/C* are closer associated with risk of death than *E, C*, and *A* directly, we fitted a Cox model on these transformed covariates. Table 2 shows the coefficients (coef), hazard ratios (HR), and p-values for each of the risk factors. (Where *A* is the age, *E* is the ejection fraction, and *C* is serum creatinine.)

**Table 2:**
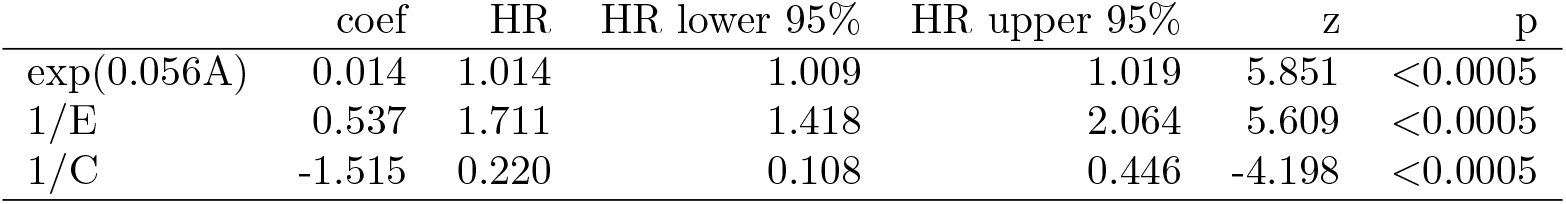
Significance of mathematically transformed covariates in the Cox model.

The log-likelihood of the model was found to be 83.8, and the C-index was 0.75. The model’s discrimination ability was also tested with a ROC curve, and the AUC was found to be 0.82 at 285 days, meaning that the model could correctly predict death within 285 days for 82% of the patients.

### Cox model with unmodified covariates

The comparison model was fitted on the same three covariates used above, but in untransformed form. Table 3 shows the coefficients (coef), hazard ratios (HR), and p-values for each of the risk factors for this comparison model.

**Table 3:** Significance of covariates without transformation in the Cox model.

|  | coef | HR | HR lower 95% | HR upper 95% | z | p |
| --- | --- | --- | --- | --- | --- | --- |
| A | 0.044 | 1.045 | 1.027 | 1.064 | 4.934 | <0.0005 |
| E | -0.049 | 0.952 | 0.933 | 0.971 | -4.803 | <0.0005 |
| C | 0.358 | 1.430 | 1.251 | 1.635 | 5.244 | <0.0005 |

The log-likelihood of the model was found to be 66.5, and the C-index was 0.72. The model’s discrimination ability with AUC was found to be 0.78 at 285 days, meaning that the model could correctly predict death within 285 days for 78% of the patients.

### Comparing the two models

Table 4 shows a comparison of all four performance metrics for the two Cox models on the three covariates (ejection fraction, serum creatinine, and age) with and without the mathematical transformations.

**Table 4:** Comparison of performance metrics.

|  | Transformed covariates | Untransformed covariates |
| --- | --- | --- |
| C-index | 0.75 | 0.72 |
| AUC (285 days) | 0.82 | 0.78 |
| Log-likelihood | 83.8 | 66.5 |
| Partial AIC | 941 | 958 |

The mathematical transformations improved the Cox model on all metrics without adding additional covariates or any other information.

## Discussion

### Beyond the linear assumption in Cox models

Cox models are a powerful tool for survival regression. They are conceptually simple, easy to interpret, and can often accurately model the survival probability over time–even given censored data. These properties have made Cox models one of the most popular methods for survival modelling in health science.

As described above, a limitation of Cox models is that each risk factor is modelled to affect risk independent of the origin. This study has shown than using mathematically transformed covariates can overcome this limitation.

For example: with the mathematical transformation 1*/E*, the hazard ratio after a unit change in ejection fraction is:

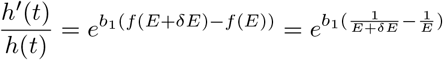

which is clearly no longer independent of the origin of E.

### Interpretability

One of the advantages of symbolic regression over other machine learning methods is that the models are easier to interpret. This is true when the full model is expressed as a symbolic relation, and it is also true when a symbolic transformation is combined with another interpretable model such as the Cox model.

For example, the fact that the transformation 1*/E* is a better risk factor than *E* directly has at least two possible interpretations:

- One is simply that the lower the ejection fraction, the larger the additional hazard given an additional reduction, i.e. a change from 0.3 to 0.29 is more dangerous than a change from 0.6 to 0.59
- Another interpretation can be seen by observing that if a fraction *E* is pumped out of the heart in every heartbeat, the average number of heart-beats that a unit of blood stays in the heart before it is pumped out will be 1*/E*, although certainly with large variation across the cavity of the ventricles.

The mathematical transformation 1*/E* of ejection fraction means that the Cox model is based on a covariate which is the average number of heartbeats blood stays in the heart.

## Conclusions

In this study, we demonstrated that combining symbolic regression with survival regression models improves the predictive power without sacrificing the interpretability that comes from a clear mathematical formulation of the model.

Other machine learning methods exist that may provide the same—or perhaps better—performance improvements than what can be achieved with the combination approach used here, but the price will typically be that the resulting model is black-box, or at least difficult to interpret.

In this study, we focused on predicting heart failure deaths, which is important in its own right, but the perspective of our research is that mathematical transformations based on symbolic regression may be applied to any survival model with improved predictive accuracy and new insights as a result.

## Data Availability

All data analysed in this study are included in the original published study by Ahmad et al. It can be directly accessed from: https://doi.org/10.1371/journal.pone.0181001.s001 (CSV)
Code to reproduce this study is publicly available under the BSD 3-Clause License at: https://github.com/wilstrup/sr-heartfailure
The code needs access to a QLattice to run. Abzu grants access to QLattices for free for research purposes. If you need access, please contact the author or the company.

https://github.com/wilstrup/sr-heartfailure

## Acknowledgements

The authors would like to thank the following for their extensive help in reviewing the article and their many suggestions for clarifications and improvements: From Abzu: Jaan Kasak, Meera Machado, and Elyse Sims. From Aalborg University: Martin Siemienski Andersen. From Zealand University Hospital: Helene Søgaard Andersen.

## Availability of data and materials

All data analysed in this study are included in the original published study by Ahmad et al[3, 2]. It can be directly accessed from: https://doi.org/10.1371/journal.pone.0181001.s001 (CSV)

Code to reproduce this study is publicly available under the BSD 3-Clause License at: https://github.com/wilstrup/sr-heartfailure

The code needs access to a QLattice to run. Abzu grants access to QLattices for free for research purposes. If you need access, please contact the author or the company.

## Ethics approval and consent to participate

The original study containing the data used in this study was approved by the Institutional Review Board of Government College University (Faisalabad, Pakistan). It states that the Helsinki Declaration were followed.[3]

## Competing interests

CW and CC are employees of Abzu, which is the company developing and the QLattice technology. CW is also co-founder of the company and the inventor of the patent-pending QLattice technology.

## Notes

### Funding Statement

No external funding received

### Author Declarations

The original study containing the data used in this study was approved by the Institutional Review Board of Government College University (Faisalabad, Pakistan). It states that the Helsinki Declaration were followed.

